# Short-term reproducibility and comparative screening performance of paired cervical cancer HPV DNA tests among women in Senegal

**DOI:** 10.64898/2026.08.18.26360727

**Authors:** Cirilus O. Osongo, Selly Ba, Marie Pierre Sy, Qinghua Feng, John Lin, Geoffrey S. Gottlieb, Papa Salif Sow, Nancy B. Kiviat, Christine J. McGrath, Stephen E. Hawes, the UW-Senegal Research Collaboration

**Affiliations:** Department of Global Health, University of Washington, Seattle, Washington, USA; Service des Maladies Infectieuses Centre Hospitalier National Universitaire (CHNU) de Fann, Dakar, Sénégal; Department of Pathology, School of Medicine, University of Washington, Seattle, Washington, USA; Department of Epidemiology, University of Washington, Seattle, Washington, USA; Center for Emerging & Re-Emerging Infectious Diseases (CERID), Division of Allergy and Infectious Diseases, Department of Medicine, School of Medicine, University of Washington, Seattle, Washington, USA

## Abstract

Cervical cancer remains a major public health challenge in sub-Saharan Africa, where access to effective screening programs remains limited. Human papillomavirus (HPV) DNA testing has emerged as a highly sensitive screening strategy for cervical precancer and cancer, although less is known about the short-term reproducibility of repeat HPV testing in high-burden settings.

This analytic observational study used secondary data from two Senegalese cohort studies conducted between 1998 and 2006 to evaluate the reproducibility and screening performance of paired cervical swab HPV DNA tests collected within 119 days of one another among 768 women. Agreement between the first and second swab HPV DNA tests was evaluated using percent agreement and Cohen’s kappa (κ) for overall high-risk HPV (hrHPV), low-risk HPV, and genotype-specific detection. Screening performance analyses compared four paired testing strategies, and exploratory logistic regression analyses examined factors associated with discordant paired hrHPV results. Reproducibility for any hrHPV detection was substantial (κ = 0.74, 95% CI: 0.69–0.79), with almost perfect agreement observed for HPV16 (κ = 0.85, 95% CI: 0.79–0.91). Agreement remained substantial across age, HIV status, education level, marital status, lifetime number of sexual partners, parity, contraceptive use, and cervical disease categories. Discordance was more likely when samples were collected 30–59 days apart than within 0–29 days and was less common among women with CIN2+, ICC, or HIV infection. Compared with a single swab strategy, classifying either swab as positive increased sensitivity for detection of both cervical intraepithelial neoplasia grade 2 or higher (CIN2+) and invasive cervical cancer (ICC) by approximately 7–8%.

This study demonstrates that paired cervical hrHPV DNA testing has substantial short-term reproducibility among women in Senegal, particularly for carcinogenic HPV types associated with cervical cancer. The findings support single hrHPV DNA testing as a reliable screening strategy in this high-burden setting while highlighting the importance of cautious interpretation of discordant repeat results, particularly among women without high-grade cervical disease.

## Introduction

Infection with oncogenic human papillomavirus (HPV) is a necessary cause of cervical cancer, which remains a leading cause of cancer-related morbidity and mortality among women worldwide, with the highest burden in sub-Saharan Africa [1,2]. Despite advances in HPV vaccination and screening, cervical cancer continues to disproportionately affect women in low- and middle-income countries (LMICs), where access to HPV vaccination, effective screening and treatment remains limited [3].

In response to the limitations of cytology-based screening, including low sensitivity, reliance on repeated testing, and substantial infrastructure requirements, HPV DNA testing has emerged as a highly sensitive method for detecting cervical precancer and cancer [4,5]. Randomized trials and meta-analyses have demonstrated greater sensitivity of HPV-based screening for high-grade cervical lesions compared with cytology and support longer screening intervals following a negative test [4,5]. Accordingly, the World Health Organization (WHO) now recommends HPV DNA testing as the preferred primary screening strategy for cervical cancer prevention, particularly in LMICs [6].

The public health utility of HPV DNA testing depends not only on disease detection but also on the reliability and reproducibility of HPV detection when testing is repeated. In clinical and programmatic settings, repeat testing may occur within weeks to months because of follow-up procedures, implementation practices, or logistical factors. The increasing adoption of self-collected samples and anticipated expansion of point-of-care and home-based molecular HPV testing further underscore the importance of understanding short-term test reproducibility [7]. Discordant results between closely timed tests may complicate clinical decision-making and patient counseling if the sources and implications of this variability are not well understood.

Experience with other molecular diagnostic tests demonstrates that closely timed results can fluctuate because of sampling variability, assay limits of detection, and short-term changes in detectable pathogen burden rather than true changes in infection status [8,9]. HPV DNA detection may similarly be influenced by variability in exfoliated cervical cell collection, specimen handling, assay performance, and viral load near the assay’s limit of detection [10,11]. Importantly, short-term changes in HPV detectability do not necessarily reflect true new acquisition or clearance of infection, which typically occurs over months to years [12]. Studies comparing clinician-collected and self-collected HPV samples have demonstrated generally high but imperfect agreement, further suggesting that sampling method and viral load may influence HPV detection [7,13]. However, these studies primarily evaluate differences between collection modalities rather than reproducibility between sequential clinician-collected samples obtained within a narrow time window.

Relatively few studies have explicitly examined short-term reproducibility of HPV DNA testing using paired cervical samples collected within days to weeks, particularly in African populations. This is an important evidence gap because HPV prevalence is higher in sub-Saharan Africa, genotype distributions differ from those in high-income settings, and conditions such as HIV infection may influence HPV viral load, persistence, and detectability [11,12,14–16]. Evaluating whether agreement varies by HIV status and other participant characteristics is therefore important for understanding test performance in populations at elevated risk of cervical cancer.

The clinical relevance of reproducibility must also be considered in relation to underlying cervical disease. Histology remains the gold standard for cervical cancer diagnosis, while the specificity of HPV testing varies by age, HPV genotype, and disease status [6,16,17]. Evaluating agreement and screening performance across the cervical disease spectrum can therefore provide insight into whether reproducibility differs among women with and without clinically significant disease and whether repeat testing provides incremental benefit beyond a single HPV test.

The paired cervical HPV samples used in this study were collected within a maximum of 119 days. This design was intended to evaluate short-term test reproducibility and concordance rather than HPV persistence or clearance, which require longer longitudinal follow-up and to reduce the likelihood that differences in HPV detection reflected longer-term changes in infection status rather than short-term variability.

Using paired HPV DNA test results from completed research studies in Senegal [18–21], the objectives of this study were to: **(1)** evaluate the short-term reproducibility of paired cervical HPV DNA testing at the overall and genotype-specific levels; **(2)** assess disease-specific agreement and the comparative screening performance of paired testing strategies for detecting cervical precancer and invasive cervical cancer; and **(3)** explore factors associated with discordant paired high-risk HPV (hrHPV) results. By evaluating these dimensions of HPV test performance in a high-burden, resource-limited setting, this study aimed to inform the interpretation of single and repeat HPV testing and the implementation of HPV-based cervical cancer screening strategies in sub-Saharan Africa and similar settings.

## Methods

### Study design and population

This study was a secondary analysis of deidentified data from two completed studies conducted in Senegal between 1998 and 2006: the EpiBio Study and the New Approaches Study. Both parent studies investigated HPV infection, cervical precancer and cancer, and related epidemiologic and clinical factors among women attending clinics in Dakar, Senegal. Published reports provide additional details on recruitment, laboratory procedures, and cervical disease findings [12,18–21].

The EpiBio Study examined the epidemiology and biology of cervical neoplasia and cancer among women aged 35 years and older recruited from Pikine, a general outpatient clinic, and Hôpital Aristide Le Dantec, a tertiary referral hospital for women with suspected cervical cancer. Of 3,354 women enrolled, between February 18, 1998, and July 26, 2003, those with HPV infection, cervical neoplasia, or suspected high grade cervical neoplasia or cervical cancer were invited for a second visit at Hôpital Aristide Le Dantec for additional evaluation and biopsy collection. The present analysis included women from this subgroup with paired cervical HPV DNA samples collected at closely timed visits.

The New Approaches Study evaluated novel biomarkers and alternative cervical cancer screening approaches among women aged 21 years and older recruited from Service des Maladies Infectieuses et Tropicales (SMIT), Centre Hospitalier National Universitaire de Fann -Fann, a national referral center for infectious disease and HIV care, and Hôpital Aristide Le Dantec. Of 1,140 women enrolled between March 12, 2003 and December 19, 2006, those undergoing additional evaluation and biopsy at a second visit were enriched for HPV positivity and cervical disease.

Although the parent studies differed in enrollment period, minimum age, and clinic setting, they used comparable questionnaire instruments, HPV testing procedures, and cytologic and histologic classifications, allowing harmonization for pooled analyses. The EpiBio cohort contributed 94% of the analytic sample.

Women were eligible if they were aged 21 years or older; had valid HPV DNA results from two cervical samples; had collection dates available for both specimens; and had samples collected within 119 days of one another. The 119-day restriction was selected to evaluate short-term reproducibility while limiting the likelihood that differences reflected longer-term HPV acquisition or clearance. Women were excluded if either HPV result was missing or invalid, specimen dates were unavailable, or cervical disease data could not be linked. The final analytic sample included 768 women recruited from Pikine, Le Dantec, and SMIT, CHNU Fann [18–21].

### Data collection and laboratory procedures

The parent studies used standardized interviews, pelvic examinations, cytology, histology, and HPV laboratory testing procedures [18–21]. Cervical specimens were collected by trained clinicians during pelvic examination using a cervical brush preserved in PreservCyt (Cytyc Corporation, Marlborough, Massachusetts, USA). Additional specimens were collected using a Dacron swab preserved in Specimen Transport Medium (Digene Corporation, Silver Spring, Maryland, USA), stored at −20°C, and shipped to Seattle for testing. Standardized collection, transport, storage, and processing protocols were used across study sites [12,18]. HPV DNA detection and genotyping were performed using polymerase chain reaction–based assays capable of detecting a broad range of HPV types. In addition to type-specific probes, the assays included a generic probe that detected HPV DNA from genotypes not individually characterized by the type-specific panel. These methods, including the Roche Linear Array platform and related genotyping assays, have been used previously in Senegalese cohorts to evaluate type-specific HPV prevalence, persistence, and clearance [19]. Quality-control procedures included standardized specimen handling, laboratory controls, duplicate testing where indicated, and verification of assay results [20].

Between 1998 and June 2003, HPV detection and typing analyses were performed via a PCR-based reverse-line strip test method (Roche Molecular Systems, Alameda, CA) with probes for HPV types 6, 11, 16, 18, 26, 31, 33, 35, 39, 40, 42, 45, 51, 52, 53, 54, 55, 56, 57, 58, 59, 66, 68, 73, 82, 83, and 84 as previously described [19]. A portion of denatured PCR products was used for line blot hybridization using Amplicon Strip Detection Reagent Kits according to the protocol recommended by the manufacturer (Roche Molecular Systems Inc., Alameda, CA). The strip contains 29 lines plus one reference ink line, detecting 27 individual HPV genotypes and 2 concentrations of the β-globin control in a single reaction. Two bovine serum albumin conjugated probes per HPV type, corresponding to each of two hypervariable regions within the MY09-11 amplicon, were deposited in a single line for each of HPV types 16, 18, 26, 31, 33, 35, 39, 42, 45, 51-59, 66, 68, 73, 82, 83 and 84. HPV types 6, 11, 40, and the β-globin controls had a single probe deposited per line.

A second PCR amplification was carried out under the same conditions except for the use of the unmodified primers. PCR products were probed with a biotin-labeled generic HPV probe. Samples hybridizing with the generic probe but not with any of the type-specific probes were considered to contain unclassified HPV. Samples that were negative for HPV DNA and yielded no β-globin gene amplification products were considered insufficient for PCR assay. In the original protocol, isolation and purification of HPV DNA took place using proteinase K digestion followed by ethanol precipitation. Beginning in October 2001, HPV DNA isolation and purification was conducted using a QIAamp DNA Mini kit (Qiagen Inc, Valencia, CA).

Beginning in June 2003, HPV detection and typing were conducted using a liquid bead microarray (LBMA) assay[20]. Sequences and probes were utilized as described above, with two probes used for HPV types 16, 18, 31, 33, 35, 39, 42, 45, 51, 52, 53, 54, 55, 58, 59, 61, 62, 66, 67, 68, 69, 71, 72, 73, 81, 82, 83, IS39 (a subtype of HPV 82), and CP6108 (also known as HPV 89), while a single probe was used for HPV types 6, 11, 26, 40, 56, 64, and 70. However, this LBMA assay uses Luminex technology for HPV genotyping, using bead sets (MiraiBio, Alameda CA) to detect 37 HPV types, and is based on the MY09-MY11-HMB01 PCR system.

ThinPrep cervical smears were evaluated by a cytotechnologist and pathologist and classified according to the Bethesda System [16,17] as normal, including reactive cellular changes; atypical squamous cells of undetermined significance (ASCUS); low-grade squamous intraepithelial lesion (LSIL); high-grade squamous intraepithelial lesion (HSIL); or squamous cell carcinoma. Biopsy specimens were evaluated by pathologists and classified as normal, cervical intraepithelial neoplasia grade 1 (CIN1), grade 2 (CIN2), grade 3 (CIN3), carcinoma in situ (CIS), or invasive cervical cancer (ICC).

Structured interviews conducted at the initial screening or enrollment visit collected demographic, behavioral, and reproductive information, including marital status, education, parity, contraceptive use, smoking, and lifetime number of sexual partners. HIV status was determined using study-specific serologic protocols that distinguished HIV-1, HIV-2, and dual HIV-1/HIV-2 infections [12,18–21].

### Study Measures

#### Participant characteristics and factors of interest

Participant characteristics included age, study site, HIV status, ethnic group, education, marital status, lifetime number of sexual partners, contraceptive use, parity, smoking, and time between paired samples.

Age was analyzed continuously and categorized as 21–49 or ≥50 years. Study site was classified as Pikine, Le Dantec, or CHNU Fann. HIV status was categorized as not living with HIV or living with HIV; the latter included HIV-1, HIV-2, and dual HIV-1/HIV-2 infection. Ethnicity was categorized as Wolof, Pular, or other, and education as none, primary, or secondary and above.

Marital status was categorized as married-polygamous, married-monogamous, formerly married, or never married. Among women in polygamous marriages, the number of co-wives was categorized as one versus two or more and examined descriptively.

Lifetime sexual partners were categorized as fewer than two versus two or more. Current contraceptive use was classified as none, hormonal, or non-hormonal. Parity was categorized as 0, 1–4, or ≥5 births, and smoking status as never or current smoker. Time between Swab 1 (first HPV DNA sample) and Swab 2 (second HPV DNA sample) was calculated in days and grouped as 0–29, 30–59, 60–89, or 90–119 days. Regression reference groups were selected *a priori* based on sample size, representativeness, and clinical interpretability.

#### Cervical disease status

Cervical disease status was defined using histology when available and cytology otherwise.

For descriptive and disease-stratified agreement analyses, disease status was harmonized into six mutually exclusive categories: normal/ASCUS, LSIL/CIN1, HSIL/CIN2+, ICC, other diagnosis, and no diagnosis available. Normal/ASCUS included normal cytology, reactive changes, and ASCUS. LSIL/CIN1 included low-grade cytologic abnormalities or histologic CIN1. HSIL/CIN2+ included HSIL or histologic CIN2, CIN3, or CIS. ICC was defined as invasive cervical carcinoma.

For screening performance and regression analyses, disease status was collapsed into three clinically relevant categories: ≤CIN1, CIN2+, and ICC. The ≤CIN1 category included normal/ASCUS and LSIL/CIN1. CIN2+ included CIN2, CIN3, and CIS but excluded ICC, which was analyzed separately.

#### HPV outcome definitions

The primary HPV outcome was detection of any high-risk HPV (hrHPV), defined according to the International Agency for Research on Cancer classification and restricted to Group 1 carcinogenic types: HPV 16, 18, 31, 33, 35, 39, 45, 51, 52, 56, 58, and 59 [21,22].

Secondary outcomes included any HPV, any low-risk HPV, and low-risk HPV only. Any HPV was defined as HPV DNA detected by either a type-specific or generic probe, regardless of oncogenic classification. This category included high-risk, low-risk, and untyped HPV-positive samples. Untyped samples were positive by the generic probe but negative for all type-specific probes.

Any low-risk HPV was defined as detection of one or more low-risk genotypes, regardless of concurrent hrHPV detection. Low-risk HPV only was defined as detection of one or more low-risk genotypes in the absence of concurrent hrHPV. Untyped infections were included in any HPV analyses but were not classified as either high-risk or low-risk HPV.

Agreement was evaluated at both grouped and genotype-specific levels. For grouped outcomes, concordance was based on positivity within the HPV category rather than persistence of the identical genotype. For example, HPV16 detected at Swab 1 and HPV18 detected at Swab 2 was classified as concordant positive for the any hrHPV analysis. Genotype-specific agreement was evaluated separately for individual hrHPV types.

For each outcome, paired results were classified as concordant positive (+/+), concordant negative (−/−), positive at Swab 1 only (+/−), or positive at Swab 2 only (−/+).

### Statistical Analysis

#### Descriptive analyses

Continuous variables with approximately symmetric distributions were summarized using the mean and standard deviation (SD), whereas variables with skewed or ordinal distributions were summarized using the median and interquartile range (IQR).

#### Objective1: Short-term agreement and reproducibility of paired HPV DNA tests

Agreement between Swab 1 and Swab 2 was evaluated for any hrHPV, any HPV, any low-risk HPV, low-risk HPV only, and individual hrHPV genotypes. Paired results were summarized as +/+, −/−, +/−, and −/+. Agreement was quantified using percent agreement and unweighted Cohen’s kappa (κ). Percent agreement was calculated as the proportion of paired observations with concordant results. Cohen’s kappa was the principal measure of reproducibility because it accounts for agreement expected by chance.

Kappa values greater than 0.80 were interpreted as almost perfect agreement, values from 0.60 to 0.80 as substantial agreement, and values from 0.40 to 0.59 as moderate agreement [23]. Values below these thresholds were interpreted as reflecting fair to poor agreement. Ninety-five percent confidence intervals for κ were estimated using nonparametric percentile bootstrap resampling with a fixed random seed of 123: 2,000 resamples for overall and genotype-specific estimates and 1,000 resamples for stratified and disease-specific estimates.

Agreement estimates were presented overall and stratified by study site, sampling interval, HIV status, age, ethnicity, education, marital status, co-wife status, lifetime sexual partners, contraceptive use, parity, smoking, and cervical disease category. These analyses were descriptive and were not intended as formal tests of interaction or differences between strata.

#### Objective 2: Disease-specific agreement and comparative screening performance

Disease-specific agreement was evaluated among women classified as normal/ASCUS, LSIL/CIN1, HSIL/CIN2+, or ICC. Women with other diagnoses or no diagnosis available were excluded from these analyses. Within each disease category, paired hrHPV results were summarized as +/+, −/−, +/−, and −/+, and percent agreement and unweighted Cohen’s kappa with 95% confidence intervals were estimated.

Comparative screening performance was evaluated for two disease endpoints: CIN2+, excluding ICC, and ICC. Women with ≤CIN1 served as the disease-negative comparison group. Four hrHPV testing strategies were assessed: Swab 1 positive, Swab 2 positive, either swab positive, and both swabs positive.

The either-swab-positive strategy evaluated whether repeat testing increased sensitivity by identifying women missed by the first swab, whereas the both-swabs-positive strategy evaluated whether requiring repeat positivity increased specificity. Sensitivity was calculated as the proportion of women with the disease endpoint who tested positive, and specificity as the proportion of women with ≤CIN1 who tested negative. Exact binomial 95% confidence intervals were calculated for all estimates. These analyses were descriptive and were not designed to formally compare diagnostic strategies through hypothesis testing.

#### Objective 3: Factors associated with discordant paired hrHPV

Exploratory logistic regression analyses examined demographic, behavioral, and clinical factors associated with discordant paired hrHPV results. Analyses were restricted to women with at least one positive hrHPV test. Women with concordant negative results were excluded because the comparison of interest was hrHPV detection at one visit only (+/− or −/+) versus detection at both visits (+/+).

Predictors included sampling interval, HIV status, cervical disease status, age, ethnicity, education, marital status, lifetime sexual partners, contraceptive use, and parity. Crude odds ratios and 95% confidence intervals were estimated using separate bivariable logistic regression models. All available observations were used for each predictor, so crude-model sample sizes varied according to missingness.

Adjusted odds ratios and 95% confidence intervals were estimated using a multivariable logistic regression model restricted to participants with complete data for all included covariates. Smoking was excluded because of sparse observations and concerns about model stability. Study site was excluded because clinic assignment reflected referral pathways and clinical populations, including referral for cervical cancer evaluation and HIV-related care, and was considered potentially intermediate to associations involving cervical disease and HIV status.

Because the regression analyses included a modest number of discordant outcomes and several sparse strata, they were considered exploratory. Interpretation emphasized the magnitude and precision of associations rather than statistical significance alone.

All inferential tests were two-sided. Analyses were performed in R version 4.5.1 (R Foundation for Statistical Computing, Vienna, Austria).

#### Missing data

Participants with missing or invalid paired HPV results or missing specimen dates were excluded during cohort assembly. For descriptive, agreement, and screening performance analyses, denominators reflected participants with non-missing data required for each analysis.

Crude regression models used all available observations for each predictor, whereas the adjusted model used complete cases for all included covariates. Missingness was low for most variables, and paired HPV outcomes were complete.

#### Ethical considerations

This study used deidentified secondary data and was classified as non-human subjects research. All participants in the parent studies provided informed consent for the collection and future use of their data under UW IRB and Senegal EC approval. The present analysis involved no direct participant contact and no access to personally identifiable information.

## Results

### Study population characteristics

A total of 768 women were included in the analytic sample (Table 1). The mean age was 45.1 years (SD, 8.83), and 73.4% were aged 21–49 years. Most participants were enrolled in the Epi/Bio Study (94.1%) and recruited from Pikine (66.4%). Nearly half (49.3%) were in polygamous marriages, 63.4% had no formal education, and 61.2% had five or more births. Overall, 9.1% of participants were living with HIV. Cervical disease status ranged from normal/ASCUS (57.6%) to invasive cervical cancer (19.4%). The median interval between paired cervical samples was 32 days (IQR, 59), with 47.0% collected within 0–29 days.

**Table 1.** Characteristics of women included in the analysis of paired cervical HPV DNA test results in Senegal (N = 768) ASCUS, atypical squamous cells of undetermined significance; CIN, cervical intraepithelial neoplasia; HIV, human immunodeficiency virus; HSIL, high-grade squamous intraepithelial lesion; IQR, interquartile range; LSIL, low-grade squamous intraepithelial lesion; SD, standard deviation.

| Characteristic | Overall |
| --- | --- |
| Age (years), mean (SD) | 45.1 (8.83) |
| Age (years), n (%) |  |
| 21- 49 | 564 (73.4) |
| ≥50 | 201 (26.2) |
| Parent study, n (%) |  |
| Epi/Bio | 723 (94.1) |
| New Approaches | 45 (5.9) |
| **Study site, n (%)** |  |
| Pikine | 510 (66.4) |
| Le Dantec | 215 (28.0) |
| CHNU Fann | 43 (5.6) |
| **Ethnic group, n (%)** |  |
| Wolof | 395 (51.4) |
| Pular | 145 (18.9) |
| Other | 228 (29.7) |
| **Lifetime sexual partners, Mean (SD)** | 1.59 (1.28) |
| Lifetime sexual partners, n (%) |  |
| 1 | 487 (63.4) |
| ≥2 | 280 (36.5) |
| **Marital status, n (%)** |  |
| Married-monogamous | 254 (33.1) |
| Married-polygamous | 379 (49.3) |
| Formerly married | 108 (14.1) |
| Never married | 15 (2.0) |
| **Number of cowives among women in polygamous marriages mean, (SD), n = 379** | 1.73 (0.97) |
| Number of co-wives among women in polygamous marriages, n (%) |  |
| 1 | 188 (49.6) |
| ≥2 | 168 (44.3) |
| Missing | 23 (6.1) |
| **Education level, n (%)** |  |
| No formal education | 487 (63.4) |
| Primary | 184 (24.0) |
| Secondary or above | 95 (12.4) |
| **Smoking status, n (%)** |  |
| Never | 754 (98.2) |
| Current | 11 (1.4) |
| **HIV status, n (%)** |  |
| Not living with HIV | 698 (90.9) |
| Living with HIV | 70 (9.1) |
| **Current contraception use, n (%)** |  |
| None | 582 (75.8) |
| Hormonal | 103 (13.4) |
| Non-hormonal | 79 (10.3) |
| **Parity, births, Median (IQR)** | 5 (4.25) |
| Parity, births, n (%) |  |
| 0 | 25 (3.3) |
| 1-4 | 261 (34.0) |
| ≥5 | 470 (61.2) |
| **Cervical disease status, n (%)** |  |
| Normal / ASCUS | 442 (57.6) |
| LSIL / CIN1 | 65 (8.5) |
| HSIL / CIN2+ | 80 (10.4) |
| Invasive cervical cancer | 149 (19.4) |
| Other diagnosis | 17 (2.2) |
| No diagnosis available | 15 (2.0) |
| **Time between paired samples, days, median (IQR)** | **32 (59)** |
| Time between paired samples, days, n (%) |  |
| 0-29 | 361 (47.0) |
| 30-59 | 175 (22.8) |
| 60-89 | 174 (22.7) |
| 90-119 | 58 (7.6) |

### Overall and genotype-specific agreement of paired HPV tests

Agreement between Swab 1 and Swab 2 was high, particularly for hrHPV detection (Fig 1; Table 2). Reproducibility for any hrHPV was substantial (κ = 0.74, 95% CI: 0.69–0.79). Agreement for any HPV was slightly lower but remained substantial (κ = 0.68, 95% CI: 0.63–0.73). Low-risk HPV outcomes also demonstrated substantial reproducibility, including any low-risk HPV (κ = 0.71, 95% CI: 0.63–0.78) and low-risk HPV only (κ = 0.61, 95% CI: 0.47–0.72). Genotype-specific analyses showed near perfect reproducibility for HPV16 (κ = 0.85, 95% CI: 0.79–0.91), while combined HPV16/18 detection also demonstrated substantial agreement (κ = 0.80, 95% CI: 0.74–0.86). Most other carcinogenic HPV genotypes demonstrated substantial agreement, although confidence intervals were wider for less prevalent types because of smaller numbers of positive observations.

**Figure 1.**
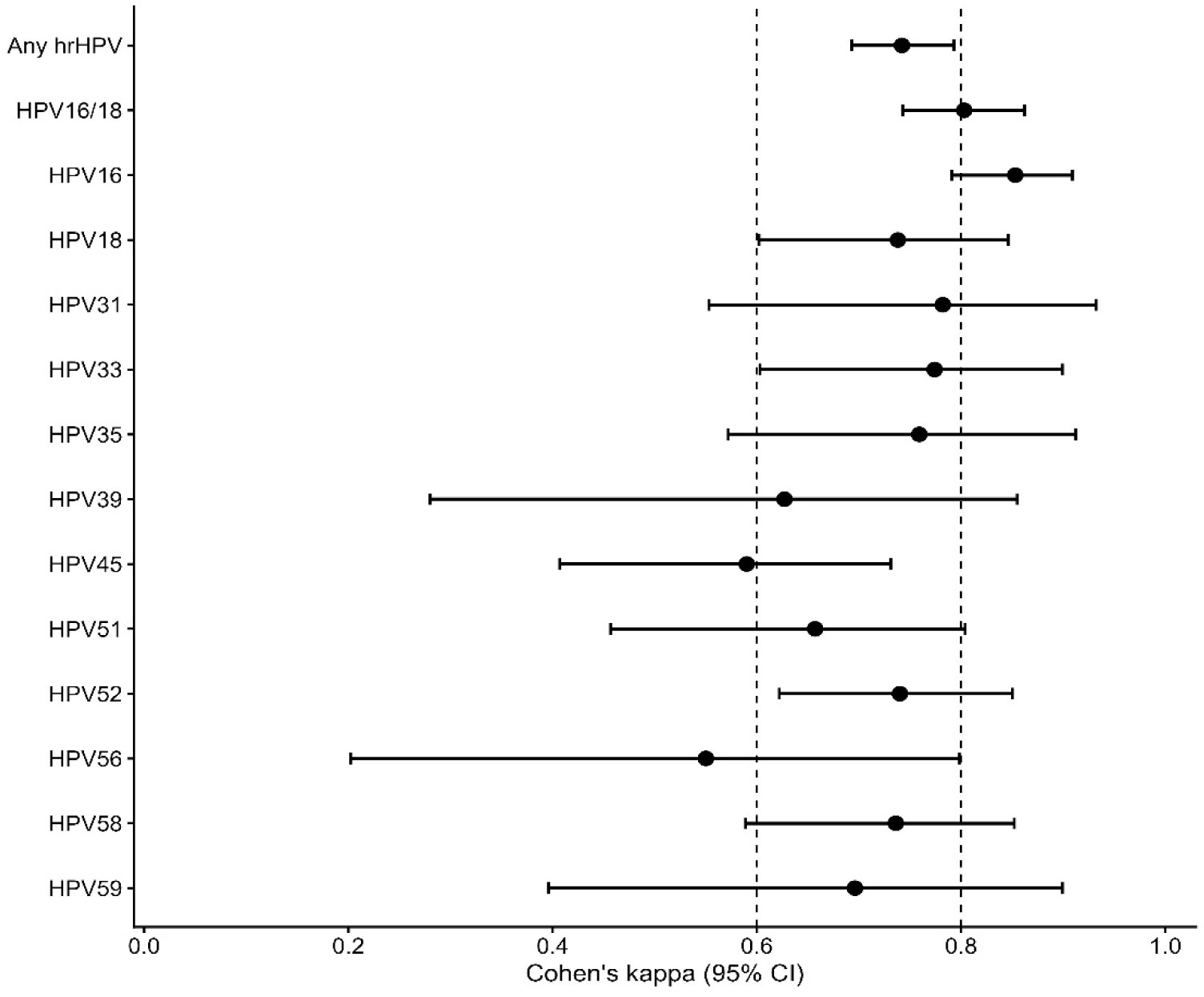
Agreement between paired cervical HPV DNA test results for overall and genotype-specific high-risk HPV detection among 768 women in Senegal. Points represent unweighted Cohen’s κ estimates and horizontal lines indicate 95% confidence intervals estimated using bootstrap resampling. Dashed vertical lines denote conventional thresholds for substantial (κ = 0.60) and almost perfect (κ = 0.80) agreement.

**Table 2.** Agreement between paired cervical HPV DNA test results for overall and high-risk genotype-specific HPV detection among women in Senegal (N = 768) CI, confidence interval; HPV, human papillomavirus; hrHPV, high-risk human papillomavirus; κ, Cohen’s kappa.

| HPV Genotype | Concordant +/+ | Concordant -/- | Discordant +/- | Discordant -/+ | Agreement (%) | Cohen's $\kappa$ (95% CI) |
| --- | --- | --- | --- | --- | --- | --- |
| Any HPV | 298 | 347 | 65 | 58 | 84.0 | 0.68 (0.63, 0.73) |
| Any hrHPV | 202 | 480 | 50 | 36 | 88.8 | 0.74 (0.69, 0.79) |
| Any Low-risk HPV | 73 | 645 | 28 | 22 | 93.5 | 0.71 (0.63, 0.78) |
| Low-risk HPV only | 26 | 711 | 15 | 16 | 96.0 | 0.61 (0.47, 0.72) |
| HPV16/18 | 104 | 623 | 22 | 19 | 94.7 | 0.80 (0.74, 0.86) |
| HPV16 | 81 | 663 | 12 | 12 | 96.9 | 0.85 (0.79, 0.91) |
| HPV18 | 27 | 723 | 10 | 8 | 97.7 | 0.74 (0.60, 0.85) |
| HPV31 | 11 | 751 | 4 | 2 | 99.2 | 0.78 (0.55, 0.93) |
| HPV33 | 16 | 743 | 5 | 4 | 98.8 | 0.77 (0.60, 0.90) |
| HPV35 | 13 | 747 | 2 | 6 | 99.0 | 0.76 (0.57, 0.91) |
| HPV39 | 6 | 755 | 4 | 3 | 99.1 | 0.63 (0.28, 0.86) |
| HPV45 | 16 | 731 | 12 | 9 | 97.3 | 0.59 (0.41, 0.73) |
| HPV51 | 14 | 740 | 5 | 9 | 98.2 | 0.66 (0.46, 0.80) |
| HPV52 | 29 | 720 | 9 | 10 | 97.5 | 0.74 (0.62, 0.85) |
| HPV56 | 5 | 755 | 5 | 3 | 99.0 | 0.55 (0.20, 0.80) |
| HPV58 | 22 | 731 | 11 | 4 | 98.0 | 0.74 (0.59, 0.85) |
| HPV59 | 7 | 755 | 3 | 3 | 99.2 | 0.70 (0.40, 0.90) |

### Agreement according to participant characteristics

Agreement for paired hrHPV detection remained generally substantial across participant subgroups (Table 3). Kappa estimates were similar across age, ethnicity, education, marital status, lifetime number of sexual partners, parity, contraceptive use, and study site, although estimates were less precise in smaller subgroups.

**Table 3.** Agreement between paired cervical high-risk HPV DNA test results according to participant characteristics among 768 women in Senegal. CI, confidence interval; HPV, human papillomavirus; hrHPV, high-risk human papillomavirus; κ, Cohen’s kappa. Number of co-wives was assessed only among women in polygamous marriages with non-missing co-wife data. Estimates from sparse strata, particularly current smokers, never-married women, nulliparous women, and participants from Fann, should be interpreted cautiously. Subgroup analyses were descriptive, and differences in κ between strata were not formally tested.

| Characteristic | Category | n | Concordant<br>+/+ | Concordant<br>-/- | Discordant<br>+/- | Discordant<br>-/+ | Agreement (%) | Cohen's $\kappa$<br>(95% CI) |
| --- | --- | --- | --- | --- | --- | --- | --- | --- |
| Overall | All participants | 768 | 202 | 480 | 50 | 36 | 88.8 | 0.74 (0.69, 0.79) |
| Study site | Pikine | 510 | 80 | 378 | 33 | 19 | 89.8 | 0.69 (0.61, 0.77) |
|  | Dantec | 215 | 92 | 95 | 15 | 13 | 87.0 | 0.74 (0.65, 0.82) |
|  | Fann | 43 | 30 | 7 | 2 | 4 | 86.0 | 0.61 (0.30, 0.86) |
| Time between samples, days | 0-29 | 361 | 117 | 208 | 16 | 20 | 90.0 | 0.79 (0.72, 0.85) |
|  | 30-59 | 175 | 25 | 127 | 15 | 8 | 86.9 | 0.60 (0.44, 0.75) |
|  | 60-89 | 174 | 46 | 108 | 16 | 4 | 88.5 | 0.74 (0.63, 0.84) |
|  | 90-119 | 58 | 14 | 37 | 3 | 4 | 87.9 | 0.71 (0.50, 0.89) |
| HIV status | Not living with HIV | 698 | 154 | 467 | 47 | 30 | 89.0 | 0.72 (0.67, 0.78) |
|  | Living with HIV | 70 | 48 | 13 | 3 | 6 | 87.1 | 0.66 (0.43, 0.85) |
| Age, years | 21-49 | 564 | 135 | 367 | 35 | 27 | 89.0 | 0.74 (0.68, 0.80) |
| | $\geq 50$ | 201 | 66 | 112 | 14 | 9 | 88.6 | 0.76 (0.66, 0.84) |
| Ethnic group | Wolof | 395 | 96 | 254 | 25 | 20 | 88.6 | 0.73 (0.65, 0.80) |
|  | Pular | 145 | 41 | 85 | 11 | 8 | 86.9 | 0.71 (0.59, 0.82) |
|  | Other | 228 | 65 | 141 | 14 | 8 | 90.4 | 0.78 (0.69, 0.86) |
| Education level | No formal education | 487 | 138 | 294 | 38 | 17 | 88.7 | 0.75 (0.68, 0.81) |
|  | Primary | 184 | 41 | 124 | 7 | 12 | 89.7 | 0.74 (0.62, 0.85) |
|  | Secondary or above | 95 | 22 | 62 | 5 | 6 | 88.4 | 0.72 (0.53, 0.86) |
| Marital status | Married-monogamous | 254 | 61 | 169 | 17 | 7 | 90.6 | 0.77 (0.68, 0.84) |
|  | Married-polygamous | 379 | 89 | 250 | 22 | 18 | 89.4 | 0.74 (0.67, 0.82) |
|  | Formerly married | 108 | 42 | 49 | 10 | 7 | 84.3 | 0.68 (0.55, 0.81) |
|  | Never married | 14 | 5 | 7 | 0 | 2 | 85.7 | 0.71 (0.29, 1.00) |
| Number of co-wives | 1 | 188 | 43 | 123 | 9 | 13 | 88.3 | 0.71 (0.61, 0.82) |
|  | ≥2 | 168 | 38 | 114 | 11 | 5 | 90.5 | 0.76 (0.65, 0.86) |
| Lifetime sexual partners | 1 | 487 | 107 | 329 | 28 | 23 | 89.5 | 0.74 (0.67, 0.80) |
|  | ≥2 | 280 | 95 | 150 | 22 | 13 | 87.5 | 0.74 (0.67, 0.82) |
| Parity, births | 0 | 25 | 6 | 15 | 2 | 2 | 84.0 | 0.63 (0.25, 0.92) |
|  | 1-4 | 261 | 81 | 155 | 15 | 10 | 90.4 | 0.79 (0.71, 0.87) |
|  | ≥5 | 470 | 111 | 305 | 32 | 22 | 88.5 | 0.72 (0.65, 0.79) |
| Current contraceptive use | None | 582 | 156 | 363 | 34 | 29 | 89.2 | 0.75 (0.69, 0.81) |
|  | Hormonal | 103 | 27 | 64 | 9 | 3 | 88.3 | 0.73 (0.58, 0.87) |
|  | Non-hormonal | 79 | 18 | 51 | 6 | 4 | 87.3 | 0.69 (0.50, 0.86) |
| Smoking status | Never | 754 | 193 | 476 | 49 | 36 | 88.7 | 0.74 (0.69, 0.79) |
|  | Current | 11 | 8 | 3 | 0 | 0 | 100.0 | 1.00 (1.00, 1.00) |

Agreement was highest among women whose samples were collected within 0–29 days (κ = 0.79, 95% CI, 0.72–0.85) and lower among those retested after 30–59 days (κ = 0.60, 95% CI, 0.44–0.75). Kappa estimates for the 60–89- and 90–119-day intervals were 0.74 (95% CI, 0.63–0.84) and 0.71 (95% CI, 0.50–0.89), respectively.

### Agreement according to cervical disease status

Agreement for paired hrHPV detection remained substantial across cervical disease categories (Table 4). Kappa estimates were 0.70 (95% CI, 0.62–0.78) among women with normal cytology or ASCUS, 0.68 (95% CI, 0.45–0.86) among women with LSIL/CIN1, 0.65 (95% CI, 0.47–0.80) among women with HSIL/CIN2+, and 0.75 (95% CI, 0.63–0.85) among women with invasive cervical cancer. Agreement (%) ranged from 82.5% among women with HSIL/CIN2+ to 90.3% among women with normal cytology or ASCUS.

**Table 4.** Agreement between paired cervical high-risk HPV DNA test results according to cervical disease status among women in Senegal (n = 736) ASCUS, atypical squamous cells of undetermined significance; CI, confidence interval; CIN, cervical intraepithelial neoplasia; HPV, human papillomavirus; hrHPV, high-risk human papillomavirus; HSIL, high-grade squamous intraepithelial lesion; κ, Cohen’s kappa; LSIL, low-grade squamous intraepithelial lesion.

| Cervical Disease Category | n | Concordant +/+ | Concordant -/- | Discordant +/- | Discordant -/+ | Agreement (%) | Cohen's $\kappa$ (95% CI) |
| --- | --- | --- | --- | --- | --- | --- | --- |
| Overall | 736 | 197 | 456 | 47 | 36 | 88.7 | 0.74 (0.70, 0.79) |
| Normal/ASCUS | 442 | 68 | 331 | 29 | 14 | 90.3 | 0.70 (0.62, 0.78) |
| LSIL/CIN1 | 65 | 13 | 44 | 3 | 5 | 87.7 | 0.68 (0.45, 0.86) |
| HSIL/CIN2+ | 80 | 38 | 28 | 8 | 6 | 82.5 | 0.65 (0.47, 0.80) |
| Invasive cervical cancer | 149 | 78 | 53 | 7 | 11 | 87.9 | 0.75 (0.63, 0.85) |

### Comparative screening performance of alternative paired testing strategies

Paired hrHPV result patterns and the screening performance of alternative testing strategies are presented in Tables 5a–5c. Among women with CIN2+, 47.5% were hrHPV positive at both swabs, while 17.5% were positive at only one swab. Among women with invasive cervical cancer, 52.3% were positive at both swabs and 12.1% were positive at only one swab. Compared with Swab 1 alone, classifying either swab as positive increased sensitivity for detecting CIN2+ and invasive cervical cancer by approximately 7–8 percentage points, while reducing specificity from 77.7% to 74.0%. Requiring both swabs to be positive increased specificity to 84.0% but reduced sensitivity to 47.5% for CIN2+ and 52.3% for invasive cervical cancer.

**Table 5a.** Distribution of paired high-risk HPV test results among women with ≤CIN1 and CIN2+ in Senegal. ASCUS, atypical squamous cells of undetermined significance; CI, confidence interval; CIN, cervical intraepithelial neoplasia; HPV, human papillomavirus; hrHPV, high-risk human papillomavirus; LSIL, low-grade squamous intraepithelial lesion.

| Outcome Category | ≤CIN1, n (%) (n = 507) | 95% CI | CIN2+, n (%) (n = 80) | 95% CI |
| --- | --- | --- | --- | --- |
| Concordant -/- | 375 (74) | 69.9, 77.7 | 28 (35.0) | 24.7, 46.5 |
| Discordant +/- | 32 (6.3) | 4.4, 8.8 | 8 (10.0) | 4.4, 18.8 |
| Discordant -/+ | 19 (3.7) | 2.3, 5.8 | 6 (7.5) | 2.8, 15.6 |
| Concordant +/+ | 81 (16.0) | 12.9, 19.5 | 38 (47.5) | 36.2, 59.0 |

**Table 5b.** Distribution of Paired High-Risk HPV Test Result Patterns Among Women With ≤CIN1 and Invasive Cervical Cancer (ICC), Senegal. ASCUS, atypical squamous cells of undetermined significance; CI, confidence interval; CIN, cervical intraepithelial neoplasia; HPV, human papillomavirus; hrHPV, high-risk human papillomavirus; LSIL, low-grade squamous intraepithelial lesion. Percentages and exact binomial 95% confidence intervals were calculated within each cervical disease category.

| Outcome Category | ≤CIN1, n (%) (n=507) | 95% CI | ICC, n (%) (n = 149) | 95% CI |
| --- | --- | --- | --- | --- |
| Concordant -/- | 375 (74.0) | 69.9, 77.7 | 53 (35.6) | 27.9, 43.8 |
| Discordant +/- | 32 (6.3) | 4.4, 8.8 | 7 (4.7) | 1.9, 9.4 |
| Discordant -/+ | 19 (3.7) | 2.3, 5.8 | 11 (7.4) | 3.7, 12.8 |
| Concordant +/+ | 81 (16.0) | 12.9, 19.5 | 78 (52.3) | 44.0, 60.6 |

**Table 5c.** Sensitivity and specificity of alternative paired high-risk HPV testing strategies for detecting CIN2+ and invasive cervical cancer among women in Senegal. ASCUS, atypical squamous cells of undetermined significance; CI, confidence interval; CIN, cervical intraepithelial neoplasia; HPV, human papillomavirus; hrHPV, high-risk human papillomavirus; LSIL, low-grade squamous intraepithelial lesion.

| Disease endpoint and testing strategy | Sensitivity, % (95% CI) | Specificity, % (95% CI) |
| --- | --- | --- |
| **CIN2+** |  |  |
| Swab 1 positive | 57.5 (45.9, 68.5) | 77.7 (73.8, 81.3) |
| Swab 2 positive | 55.0 (43.5, 66.2) | 80.3 (76.5, 83.7) |
| Either swab positive | 65.0 (53.5, 75.3) | 74.0 (69.9, 77.7) |
| Both swab positive | 47.5 (36.2, 59.0) | 84.0 (80.5, 87.1) |
| **Invasive cervical cancer** |  |  |
| Swab 1 positive | 57.0 (48.7, 65.1) | 77.7 (73.8, 81.3) |
| Swab 2 positive | 59.7 (51.4, 67.7) | 80.3 (76.5, 83.7) |
| Either swab positive | 64.4 (56.2, 72.1) | 74.0 (69.9, 77.7) |
| Both swab positive | 52.3 (44.0, 60.6) | 84.0 (80.5, 87.1) |

### Factors associated with discordant paired hrHPV results among women with at least one positive hrHPV test

Exploratory regression analyses were restricted to 288 women with hrHPV detected on at least one of the two swabs. Of these, 86 (29.9%) had discordant paired results, defined as hrHPV detection at only one swab (+/− or −/+), and 202 (70.1%) had concordant positive results, defined as hrHPV detection at both swabs (+/+) (Table 6). Crude analyses used all available observations for each predictor, whereas the multivariable analysis was restricted to 268 women with complete data for all included covariates.

**Table 6.**
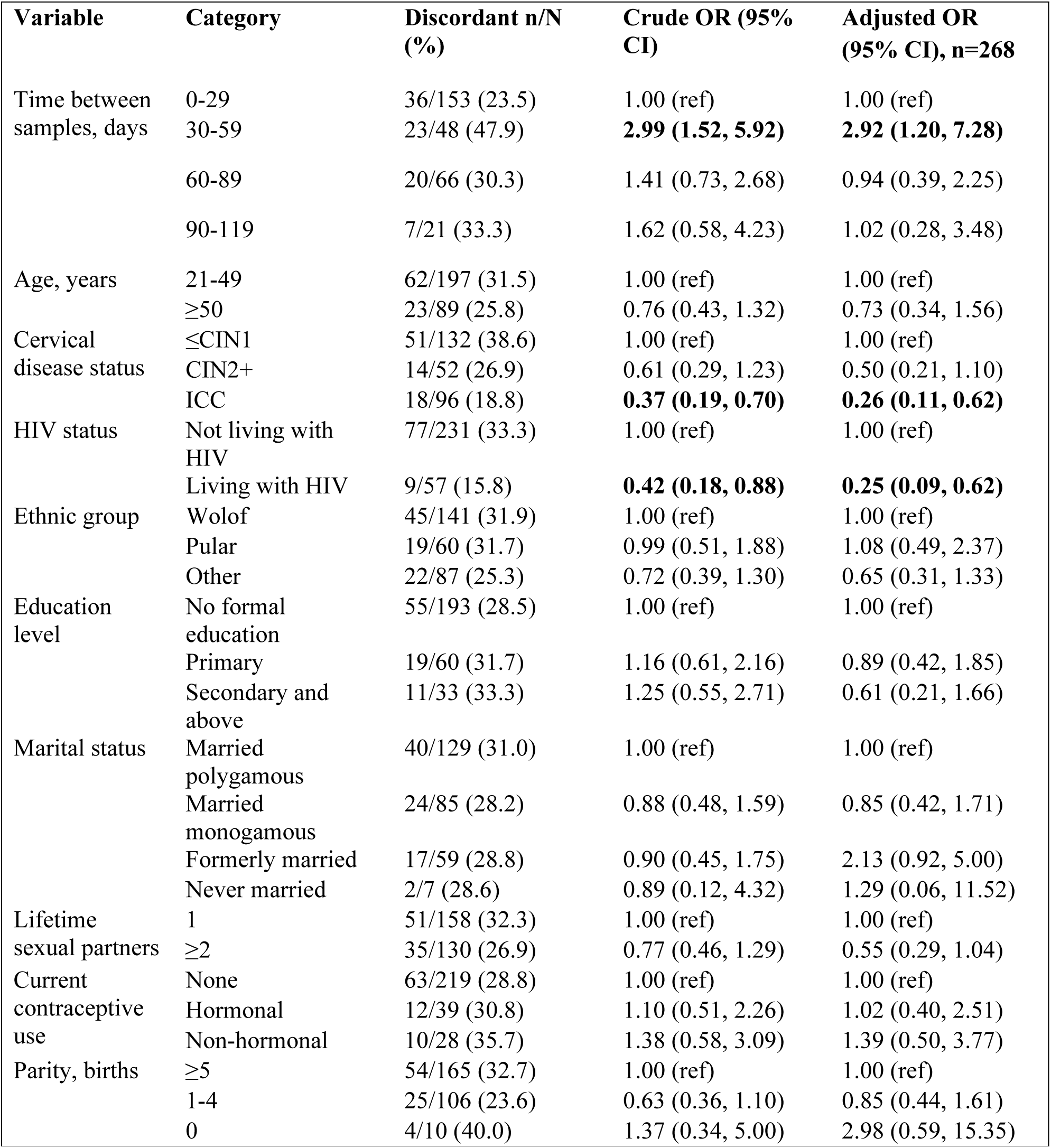
Factors Associated with discordant paired high-risk HPV test results among women with high-risk HPV detected on at least one swab in Senegal. CI, confidence interval; CIN, cervical intraepithelial neoplasia; HIV, human immunodeficiency virus; HPV, human papillomavirus; hrHPV, high-risk human papillomavirus; OR, odds ratio; ref, reference category. Study site was excluded from the final adjusted model because clinic assignment reflected underlying referral patterns and clinical populations, including referral for cervical cancer evaluation and HIV-related care, and was considered potentially intermediate to associations involving cervical disease severity and HIV status. Smoking was excluded because of sparse data and concerns about model stability. Reference categories were selected a priori based on sample size, representativeness of the study population, and clinical interpretability. Estimates from sparse strata should be interpreted cautiously.

Compared with women whose paired samples were collected 0–29 days apart, women with samples collected 30–59 days apart had higher adjusted odds of discordant results (adjusted OR = 2.92, 95% CI, 1.20–7.28). No clear increase in the odds of discordance was observed for samples collected 60–89 or 90–119 days apart.

Discordance was less common among women with more severe cervical disease. Discordant results occurred in 38.6% of women with ≤CIN1, 26.9% of women with CIN2+, and 18.8% of women with invasive cervical cancer. Compared with women with ≤CIN1, women with invasive cervical cancer had lower adjusted odds of discordant results (adjusted OR = 0.26, 95% CI, 0.11–0.62). Women with CIN2+ also had lower estimated odds of discordance, although the confidence interval included the null (adjusted OR = 0.50, 95% CI, 0.21– 1.10).

Women living with HIV had less frequent discordance than women not living with HIV (15.8% vs 33.3%) and lower adjusted odds of discordant paired hrHPV results (adjusted OR = 0.25, 95% CI, 0.09–0.62). Most other participant characteristics were not clearly associated with discordant paired hrHPV results after multivariable adjustment.

## Discussion

This study evaluated the short-term reproducibility and screening performance of paired cervical HPV DNA tests collected within 119 days among 768 women in Senegal. Paired hrHPV testing showed substantial reproducibility overall, with the strongest agreement observed for HPV16 and combined HPV16/18 detection. Agreement was generally consistent across participant subgroups and cervical disease categories. Among women with hrHPV detected on at least one swab, discordance was more frequent when samples were collected 30-59 days apart than within 0-29 days and less frequent among women with invasive cervical cancer or HIV infection. Classifying either swab as positive modestly increased sensitivity for CIN2+ and invasive cervical cancer, whereas requiring both swabs to be positive increased specificity at the expense of sensitivity. These findings support the reliability of cervical HPV DNA testing in this setting while clarifying the interpretation and potential value of closely timed repeat testing.

The substantial agreement observed for any hrHPV complements the broader evidence supporting HPV DNA testing for cervical cancer screening. HPV-based screening is more sensitive than cytology for detecting high-grade cervical precancer and provides greater protection against invasive cervical cancer in randomized trials and meta-analyses [5,24]. Ronco et al. reported greater protection against invasive cervical cancer with HPV-based screening than with cytology [5], while Koliopoulos et al. found higher sensitivity for CIN2+ and CIN3+, although with lower specificity [24]. The present findings extend this evidence by demonstrating that cervical HPV DNA testing also produces generally consistent results when repeated over a short interval in a West African population.

Agreement was particularly strong for HPV16, which demonstrated almost perfect reproducibility. This is clinically important because HPV16 is the most carcinogenic HPV genotype and accounts for the largest proportion of invasive cervical cancers worldwide [25,26]. A recent global systematic analysis estimated that HPV16 is attributable to approximately 61.7% of invasive cervical cancer cases, substantially more than any other HPV genotype [25]. Khan et al. further showed that HPV16 and HPV18 positivity predicted elevated long-term risk of cervical precancer and cervical cancer [26]. The high reproducibility of HPV16 observed in this study suggests that infections most relevant to cervical carcinogenesis are consistently detected across closely timed repeat samples, strengthening confidence in HPV DNA testing for identifying clinically important carcinogenic infections Despite high overall agreement, discordant paired results occurred in a meaningful minority of women with at least one positive hrHPV test. These results should not be interpreted automatically as true acquisition or clearance, given the short interval between samples. Natural history studies indicate that HPV acquisition, persistence, and clearance generally occur over months to years rather than within the time frame evaluated here [27–30]. Ho et al. documented that many cervicovaginal HPV infections clear over time [27], while Rodríguez et al. emphasized that persistent rather than transient infection is the principal concern for cervical cancer prevention [28]. Short-interval discordance is therefore more likely to reflect variability in cervical cell sampling, intermittent viral shedding, specimen quality, or viral load near the assay’s detection threshold.

The distinction between analytical and clinical sensitivity further informs this interpretation. Snijders et al. noted that highly sensitive HPV assays can detect low-level infections that may have limited clinical significance, whereas clinically useful testing requires balancing detection with disease prediction [29]. A positive result on one swab and a negative result on the other may therefore represent a genuine low-level infection that was inconsistently sampled or detected rather than a meaningful biologic transition. Discordance is thus an expected feature of molecular testing when viral burden is low and should be interpreted in conjunction with disease status and clinical context.

The association between sampling interval and discordance was non-linear. Agreement was highest within 0– 29 days, and women retested after 30–59 days had higher adjusted odds of discordant results, but no progressive increase was observed across longer intervals. If acquisition or clearance were the dominant explanation, discordance might be expected to rise steadily as the interval increased. The isolated increase at 30–59 days may instead reflect random variability, smaller subgroup sizes, or unmeasured clinical and programmatic factors. Short-term fluctuation near assay detection thresholds has also been observed with other molecular tests [30,31]. Although SARS-CoV-2 and HPV differ biologically, the broader principle that repeated PCR results may alternate when target concentrations are low remains relevant to interpreting closely timed HPV tests.

Women living with HIV had lower adjusted odds of discordant paired hrHPV results than women not living with HIV, indicating more consistent detection across the two samples. HIV infection is associated with higher HPV prevalence, more frequent multiple infections, altered clearance, and greater risk of cervical precancer and cancer [14,18,32–36].Studies from Senegal have also documented increased risks of high-grade cervical lesions and invasive cervical cancer among women with HIV-1 and HIV-2 infections [18,32]. Liu et al. similarly summarized the elevated burden of HPV infection and cervical disease among women living with HIV[33]. Gingles et al. found that 12 months after thermal ablation, hrHPV persistence was significantly higher among women living with HIV than women without HIV (46.0% vs. 32.0%), consistent with altered viral clearance in this population [34]. Rao et al. confirmed that women living with HIV face a six-to ten-fold increased risk of cervical cancer attributable to higher HPV prevalence and persistence, with high-risk HPV positivity of 24.2% and a notable burden of high-grade lesions detected on screening [35]. Higher HPV viral loads may contribute to more stable molecular detection; Hanisch et al. reported higher HPV16 viral load in relation to HIV infection and cervical disease in Senegal [36]. The lower discordance observed among women living with HIV is therefore consistent with a greater probability of persistent, detectable infection, although the relatively small HIV-positive subgroup warrants cautious interpretation.

Cervical disease status also helped clarify the clinical meaning of discordance. Agreement remained substantial across the disease spectrum, while discordance was less frequent among women with invasive cervical cancer and was also lower, although less precisely estimated, among women with CIN2+ than among women with ≤CIN1. Persistent carcinogenic HPV infection is central to cervical carcinogenesis, and clinically significant lesions are more likely to be associated with stable and detectable hrHPV infection [37–39]. Bosch et al. described the causal role of persistent carcinogenic HPV infection in cervical cancers [37,38], and de Sanjosé et al. demonstrated that HPV DNA is detected in most invasive cervical cancers worldwide, with HPV16 and HPV18 accounting for a large share of cases [39]. The greater consistency of repeat detection among women with more severe disease is therefore compatible with the biology of persistent carcinogenic infection.

The screening analyses illustrated the trade-off introduced by repeat testing. Classifying either swab as positive increased sensitivity for CIN2+ and invasive cervical cancer by approximately 7–8 percentage points compared with Swab 1 alone, but specificity decreased. Requiring both swabs to be positive produced the opposite pattern, increasing specificity while reducing sensitivity. The trade-off between sensitivity and specificity when combining two test results has similarly been described across other diagnostic contexts, including point-of-care proteinuria testing, and sideline concussion assessment [40,41]. Although these conditions differ biologically from HPV infection, the broader principle that requiring joint positivity across two tests improves sensitivity at the expense of specificity remains relevant to interpreting paired hrHPV results. A second test may identify women missed initially, but its value depends on whether the incremental sensitivity justifies the additional testing, follow-up, and treatment demands.

These considerations are particularly important in resource-limited settings. WHO recommends HPV DNA testing as the preferred primary screening approach where feasible because of its high sensitivity and potential to support longer screening intervals after a negative result [6]. However, screening programs must also account for laboratory capacity, follow-up systems, treatment availability, and patient costs [42]. The modest sensitivity gain observed with an either-swab-positive strategy may not justify universal repeat testing. Repeat testing may be more appropriate in selected circumstances, such as inadequate specimens, uncertain initial results, high-risk clinical groups, or programmatic workflows in which testing occurs at more than one visit. Conversely, requiring positivity on both swabs could reduce false-positive referrals but would miss additional cases and may be unsuitable where loss to follow-up is common.

This study has several strengths. It used paired cervical specimens from a well-characterized Senegalese population, allowing direct evaluation of short-term reproducibility rather than cross-sectional positivity alone. The inclusion of women across the cervical disease spectrum enabled assessment of agreement and screening performance against clinically relevant endpoints. Genotype-specific PCR-based testing permitted evaluation of individual carcinogenic HPV types, including HPV16 and HPV18. The study also addressed a practical screening question by examining whether repeat testing yields consistent results and whether a second test changes sensitivity and specificity.

Several limitations should be considered. The data were collected between 1998 and 2006, and HPV testing platforms have evolved since that period. Nevertheless, PCR-based genotyping remains central to HPV epidemiology and assay validation, and the paired-sample design remains informative for reproducibility. HrHPV detection among women with invasive cervical cancer was lower than the expected prevalence of more than 95%, which may reflect sampling variability, suboptimal storage and transport practices, elapsed time between collection and assaying, low cellular yield in advanced lesions, assay limitations, or heterogeneity in HPV DNA detectability within tumors. Because this was a secondary analysis, the interval between swabs was not randomized and may have reflected clinical or programmatic factors. Estimates were also imprecise in sparse strata, including current smokers, nulliparous women, never-married women, and women with less common genotypes. Finally, this study evaluated short-term reproducibility rather than persistence, clearance, or progression; those outcomes have been examined more directly in longitudinal analyses from the same Senegalese cohorts [18–20,32].

The study population was drawn from both general outpatient and referral settings and was enriched for HPV positivity and cervical disease. This broadened the disease spectrum available for analysis but may limit generalizability to routine population-based screening. Sensitivity and specificity estimates should therefore be interpreted in relation to the underlying recruitment design rather than assumed to apply directly to general screening populations. None of the participants had received HPV vaccination because enrollment preceded widespread vaccine implementation in Senegal and much of sub-Saharan Africa. Generalizability to vaccinated populations may consequently be limited, as vaccination can alter genotype distribution, overall carcinogenic HPV prevalence, and the relative contribution of HPV16 and HPV18 to cervical disease [43,44].

Future studies should evaluate short-term reproducibility using newer HPV platforms, self-collected samples, point-of-care assays, and vaccinated populations. Clinician-collected and self-collected samples generally show good agreement, although performance varies by collection method, assay, and viral load [45–48]. As self-sampling and decentralized screening expand, understanding reproducibility across collection strategies will become increasingly important. Longitudinal studies should also determine whether short-interval discordance predicts subsequent persistence, clearance, or disease progression and whether discordant results are concentrated among infections with low viral burden or limited clinical relevance.

In conclusion, cervical hrHPV DNA testing demonstrated substantial short-term reproducibility among women in Senegal, particularly for HPV16 and other carcinogenic types of greatest relevance to cervical cancer prevention. Repeat testing modestly increased sensitivity when either swab was considered positive but reduced specificity, indicating that routine repeat testing should be considered in relation to its programmatic costs and clinical consequences. Discordance was less frequent among women with invasive cervical cancer and HIV infection, consistent with more persistent or readily detectable infections in these groups. These findings support the reliability of HPV-based screening in high-burden settings while emphasizing that short-interval discordance should be interpreted cautiously and in the context of disease severity, viral burden, and sampling variability.

## Data Availability

The deidentified minimal dataset underlying the findings of this study and the analytic code will be made available in accordance with applicable data-governance requirements and PLOS ONE’s data availability policy. Repository and access information will be provided prior to publication.

## Acknowledgments

This study would have not been successful if it were not for the many women who accepted to participate and the research teams at the University of Washington and in Senegal for their dedication to this research.

